# Association Between a 22-feature Genomic Classifier and Biopsy Gleason Upgrade During Active Surveillance for Prostate Cancer

**DOI:** 10.1101/2021.11.22.21266727

**Authors:** Benjamin H. Press, Tashzna Jones, Olamide Olawoyin, Soum D. Lokeshwar, Syed N Rahman, Ghazal Khajir, Daniel W. Lin, Matthew R. Cooperberg, Stacy Loeb, Burcu F. Darst, Yingye Zheng, Ronald C. Chen, John S. Witte, Tyler M. Seibert, William J. Catalona, Michael S. Leapman, Preston C. Sprenkle

**Author notes:** <u>Corresponding Author:</u> Preston C. Sprenkle, MD, Associate Professor of Urology, Division Chief, Division of Urology at VA Connecticut Healthcare System, Director, Urology Research Fellowship, Director, Urologic Oncology Clinical Fellowship, Department of Urology, Yale School of Medicine, New Haven, Connecticut.

## Abstract

**Background:** Although the Decipher genomic classifier has been validated as a prognostic tool for several prostate cancer endpoints, little is known about its role in assessing risks of biopsy reclassification among patients on active surveillance, a key event that often triggers treatment

**Objective:** To evaluate the association between Decipher genomic classifier and biopsy Gleason upgrade among patients on active surveillance.

**Design, Setting, and Participants:** Retrospective cohort study among patients with low- and favorable-intermediate-risk prostate cancer on active surveillance who underwent biopsy-based Decipher testing as part of clinical care.

**Outcomes measures and statistical analysis:** Any increase in biopsy Gleason grade group (GG). We evaluated the association between Decipher score using univariable and multivariable logistic regression. We compared area under the receiver operating characteristic curve (AUC) of models comprised of baseline clinical variables with or without Decipher score.

**Results and limitations:** We identified 133 patients of median age 67.7 years and median PSA 5.6 ng/mL. At enrollment 75.9% were GG1 and 24.1 GG2. Forty-three patients experienced biopsy upgrade. On multivariable logistic regression, Decipher score was significantly associated with biopsy upgrade (OR 1.37 per 0.10 unit increase, 95% CI 1.05-1.79 p=0.02). Decipher score was associated with upgrade among patients with biopsy Grade group 1, but not Grade Group 2 disease. The discriminative ability of a clinical model (AUC 0.63, 95% CI 0.51-0.74) was improved with the integration of Decipher score (AUC 0.69, 95% CI 0.58-0.80).

**Conclusions:** The Decipher genomic classifier was associated with short-term biopsy Gleason upgrading among patients on active surveillance.

**Patient summary:** The results from this study indicate that among patients with prostate cancer undergoing active surveillance, those with higher Decipher scores were more likely to have higher-grade disease found over time. These findings indicate that the Decipher test might be useful for guiding the intensity of monitoring during active surveillance, such as more frequent biopsy for patients with higher scores.

## Introduction

Active surveillance is the recommended initial management strategy for most patients with low-grade prostate cancer and an option for selected patients with favorable-intermediate risk disease and is now adopted by the majority of eligible patients.^1^ Evidence from randomized trials and institutional cohort studies supports the long-term safety of active surveillance and its effectiveness as a strategy to avoid or defer definitive treatment.^2,3^ Nonetheless, 20-60% of patients who are initially enrolled in active surveillance ultimately experience reclassification of their disease based on changes in biopsy Gleason grade, PSA levels, or cancer volume.^4,5^ As a result, up to half of the patients undergo definitive treatment in the near term, most frequently due to Gleason upgrading.^6^ A smaller number of patients with clinically low-risk features ultimately experience clinically significant progression over time, underscoring the need for close monitoring to detect early signs of reclassification.^7^ Estimating the risk of disease reclassification during active surveillance based on standard clinical parameters is imperfect, leading to patient anxiety, avoidable treatment, and imprecision in monitoring (e.g., over or under-use of surveillance testing).^8-10^

Genomic classifiers measuring features associated with prostate cancer aggressiveness developed largely in patients with high-risk disease provide robust predictions of disease outcome, yet little is known about their role in estimating the trajectory of untreated favorable-risk prostate cancer.^11^ The Decipher classifier (GenomeDx Biosciences, Vancouver, BC, Canada), is a tissue-based platform evaluating the expression of 22 genes selected from whole-transcriptome analysis and reflect pathways involved in cellular proliferation, differentiation, immune modulation, and androgen-receptor signaling. The test has been widely validated both as a prognostic and predictive marker associated with several clinical outcomes, including adverse pathology at prostatectomy, biochemical recurrence, metastasis, and prostate cancer mortality after treatment.^12,13^ However, less information is available regarding their utility in predicting the outcome of patients being managed with active surveillance. Such information would be useful as a means to tailor the approaches to clinical management – potentially moderating surveillance protocols for those at lowest-risk and intensifying or foregoing surveillance in patients most likely to experience reclassification or disease progression.^8^

In this study, we aimed to evaluate the association between the Decipher genomic classifier and biopsy outcomes among patients with favorable-risk prostate cancer.^14^ Analytic and clinical validation of commercially available genomic tests were largely conducted using archival tissue obtained in the era before widespread use of prostate magnetic resonance imaging (MRI), an approach that significantly improves the accuracy of sampling and reduces the risk of initial misclassification.^5,15,16^ Therefore, commensurate with current clinical practice, we further sought to conduct our study among a contemporary cohort of patients managed with active surveillance following an MRI-ultrasound fusion-guided prostate biopsy.

## Materials and Methods

### Study Design and Patient Selection

We performed a retrospective cohort study of patients enrolled on active surveillance for prostate cancer who underwent Decipher testing. We identified subjects from a prospectively maintained institutional repository of patients with known or suspected prostate cancer undergoing prostate MRI and prostate biopsy at a single tertiary care center. The primary study objective was to examine the association between a patient’s baseline Decipher score (scale 0-1.0 units) and Gleason upgrading during active surveillance, defined as an increase in the Gleason grade group on subsequent biopsy. The secondary objectives were to evaluate the performance of clinical prediction models with or without the genomic classifier, and to identify a clinical threshold for the Decipher score in predicting Gleason upgrading. In addition, we evaluated the association between Gleason upgrade and the clinically reported Decipher risk groups: low (<0.45), intermediate (0.45-0.60), and high (>0.60).

Of 1,432 patients undergoing prostate MRI-ultrasound fusion biopsy, we identified 133 who elected initial active surveillance with at least one additional biopsy and underwent Decipher testing from July 2016 through November 2020. Patients with low-risk prostate cancer (Gleason score ≤ 3+3, clinical-stage T1 [cT1], PSA ≤ 10 ng/mL) and select patients with favorable intermediate-risk prostate cancer (Gleason score ≤ 3+4 with ≤ one core with Gleason pattern 4, ≤ cT2, PSA 10-20 ng/mL) detected on combined systematic and MRI-ultrasound-fusion-targeted biopsy (MRF-TB) were enrolled into the active surveillance program and included in an IRB-approved prospective data registry. The institutional surveillance protocol consisted of semi-annual PSA testing, a confirmatory prostate biopsy within one year of diagnosis, and subsequent prostate MRI and prostate biopsy on a yearly or biennial basis. Protocols for MRI and MRF-TB were conducted in a manner previously described.^17^ Genomic testing was routinely offered to patients considering active surveillance without restriction based on disease characteristics. We compiled clinical, pathology, and sociodemographic information, including prostate MRI findings and the Decipher score.

### Statistical Analysis

We compiled clinicopathologic variables, Decipher scores, and biopsy upgrade status for each patient. Categorical variables were reported as *n* (%); continuous variables were reported as the median and interquartile range (IQR). We used McNemar’s test for statistical analysis of proportions, and the Kruskal–Wallis test was used for continuous variables. We constructed multivariable logistic regression models to evaluate the association between baseline characteristics, including the Decipher score and biopsy Gleason upgrading. Variables that were significantly associated with upgrading on univariable analysis were included in the model, as well as *a priori* variables shown to be associated with Gleason upgrading in prior studies (age, PSA density, number of biopsy cores positive for cancer and prostate MRI findings). We compared the performance of a baseline clinical model with the Decipher classifier alone, and a combined model consisting of clinical parameters and Decipher score. We used Youden’s index to identify a potential threshold of Decipher score that could be clinically used to identify patients at greater risk for reclassification during active surveillance. All statistical analyses were performed using SPSS version 27 IBM SPSS Statistics, Armonk, NY, USA).

## Results

The study sample consisted of 133 patients initially managed with active surveillance who received Decipher testing. The median age at enrollment was 67.7 years (IQR 62.4 – 71.4), and the median PSA at diagnosis was 5.6 ng/mL (IQR 4.3 – 7.1), Table 1. In this cohort, 66 (49.6%) had Decipher testing performed on their initial diagnostic biopsy and 67 (50.4%) had testing on a subsequent biopsy. The biopsy Gleason grade at enrollment was GG1 for 75.9% and GG2 for 24.1%. The median interval between biopsies was 13.6 months (IQR 11.9-16.9), and the median Decipher score was 0.39 (IQR 0.25-0.48). The distribution of reported Decipher risk groupings was ‘low’ in 64.4%, ‘intermediate’ in 25.3%, and ‘high’ in 10.3% of patients. Changes in prostate MRI PI-RADS scores occurred in 41 patients (30.7%).

**Table 1.** Baseline characteristics of patients with favorable-risk prostate cancer undergoing active surveillance and receiving Decipher genomic testing.

| Variable | Value |
| --- | --- |
| **Median age in years, (IQR)** | 67.7 (62.4 – 71.4) |
| **BMI, median (IQR)** | 27.1 (25.0 – 30.5) |
| **PSA (ng/mL), median (IQR)** | 5.6 (4.3 – 7.1) |
| **Biopsy Gleason Grade Group 1, N(%)** | 101 (75.9) |
| **Biopsy Gleason Grade Group 2, N(%)** | 32 (24.1) |
| **Race/Ethnicity, N(%)** |  |
| White | 120 (90.2) |
| Black/African-American | 8 (6.0) |
| Latino | 3 (2.3) |
| Other | 2 (1.5) |
| **Prostate Volume, median (IQR)** | 46.0 (32.6 – 59.0) |
| **Decipher Score, median (IQR)** | 0.39 (0.39 – 0.48) |
| **Decipher Risk Category** |  |
| Low (<0.45) | 94 (64.4) |
| Intermediate (0.45-0.60) | 37 (25.3) |
| High (>0.60) | 15 (10.3) |
| *Abbreviations: IQR= interquartile range; BMI=body mass index;*<br>*PSA=prostate specific antigen* |  |

**Table 2.** Comparison of characteristics among patients who did and did not experience biopsy Gleason upgrading during active surveillance.

| Variable | Did Not Experience<br>Biopsy Gleason Upgrade<br>(N = 90 ) | Biopsy Gleason Upgrade<br>(N = 43 ) | P-value |
| --- | --- | --- | --- |
| Median Age (IQR) | 68.0 (62.4 – 70.9) | 66.3 (61.9 – 73.2) | 0.93 |
| Median PSA (ng/mL), (IQR) | 5.6 (4.1 – 7.0) | 5.6 (4.4 – 7.2) | 0.97 |
| Median BMI (IQR) | 26.8 (24.7 – 30.4) | 28.1 (25.3 – 30.9) | 0.20 |
| Median PSA Density (IQR) | 0.12 (0.08 - .18) | 0.12 (0.08 – 0.16) | 0.69 |
| Median Prostate Volume (IQR) | 45.0 (33.5 – 57.1) | 47.0 (31.0 – 60.0) | 0.81 |
| Decipher Risk Category, (%) |  |  | 0.27 |
| Low (<0.45) | 60 (0.67) | 24 (55.8) |  |
| Intermediate (0.45-0.60) | 23 (0.26) | 12 (27.9) |  |
| High (>0.60) | 7 (0.08) | 7 (16.3) |  |
| Median Decipher (IQR) | 0.39 (0.25 – 0.46) | 0.41 (0.32 – 0.54) | 0.06 |
| Median Systematic Positive | 2 (1 – 4) | 3 (1 – 4) | 0.53 |
| Prostate MRI PI-RADS, N (%) |  |  | 0.57 |
| 1-2 | 22 (25.0) | 8 (18.6) |  |
| 3 | 7 (7.9) | 5 (11.7) |  |
| 4 | 39 (44.4) | 20 (46.5) |  |
| 5 | 20 (22.7) | 10 (23.2) |  |
| Increase in MRI PI-RADS<br>Classification | 22 (28.2) | 10 (32.2) | 0.68 |
| Abbreviations: *IQR*= interquartile range; *BMI*=body mass index; *PSA*=prostate specific antigen |  |  |  |

**Table 3.** Multivariable logistic regression model examining factors associated with biopsy Gleason upgrade during active surveillance.

| Variable | Odds Ratio | 95% Confidence Interval |  | *p* |
| --- | --- | --- | --- | --- |
|  |  | Lower | Upper |  |
| Age | 1.02 | 0.97 | 1.07 | 0.44 |
| Baseline Prostate MRI PIRADS |  |  |  | 0.77 |
| 1 and 2 | Reference | - | - |  |
| 3 | 0.63 | 0.14 | 2.82 | 0.55 |
| 4-5 | 1.02 | 0.36 | 2.89 | 0.97 |
| PSA Density (per 0.1 unit) | 0.83 | 0.50 | 1.44 | 0.52 |
| Decipher Score (per 0.1 unit) | 1.37 | 1.05 | 1.79 | 0.02 |
| ≥3 Positive systematic biopsy cores | 2.55 | 1.04 | 6.29 | 0.04 |
| Abbreviations: PI-RADS=Prostate Imaging Reporting and Data System |  |  |  |  |

In total, 43 patients (32.3%) experienced biopsy upgrading. The median Decipher score among those who upgraded was 0.39 (IQR 0.25-0.46) as compared with 0.41 (IQR 0.32 – 0.54) among patients who were not (p=0.06). The distribution of upgrading events did not differ significantly by Decipher risk groups (28.6% [low risk], 34.3% [intermediate risk], 50.0% [high risk], p=0.27). On univariable analysis, increasing Decipher score was associated with greater odds of upgrading (OR 1.24 per 0.10 unit, 95% CI, p=0.045). When stratified by the diagnostic Gleason grade group, the Decipher score was associated with upgrading among patients with GG1, (OR 1.29 per 0.10 unit, p=0.047), but not among those with GG2 disease (p=0.41). On multivariable logistic regression analysis, Decipher score remained significantly associated with the odds of biopsy upgrading (OR 1.37 per 0.10 units, 95% CI p=0.02).

The baseline clinical model showed modest discrimination of biopsy upgrade (AUC 0.63, 95% CI 0.51-0.74). The AUC for Decipher alone was 0.60 (95% 0.49-0.70). A combined model including Decipher score and clinical variables improved the AUC to 0.69 (95% CI 0.58-0.80). Figure 1. A Decipher cutoff of 0.475 maximized sensitivity and specificity for the prediction of biopsy upgrade while on AS. At a dichotomous threshold of 0.475, sensitivity and specificity for biopsy upgrading were 41.9% and 78.9%, respectively, and showed modest discrimination of biopsy upgrade (AUC = 0.60, 95%CI 0.52-0.69). Among patients with Decipher scores <0.475 versus ≥0.475, the incidence of biopsy upgrading was 26.0% vs 48.6%, respectively, p=0.02). On univariable analysis, Decipher scores greater than or equal to 0.475 were associated with increased odds of biopsy upgrade (OR 2.69, p = 0.01, 95%CI 1.22-5.92). On multivariable logistic regression analysis, Decipher scores above the cutoff of 0.475 were independently associated with odds of biopsy upgrade (OR 3.71, 95% CI 1.45-9.50, p=0.01).

**Figure 1.**
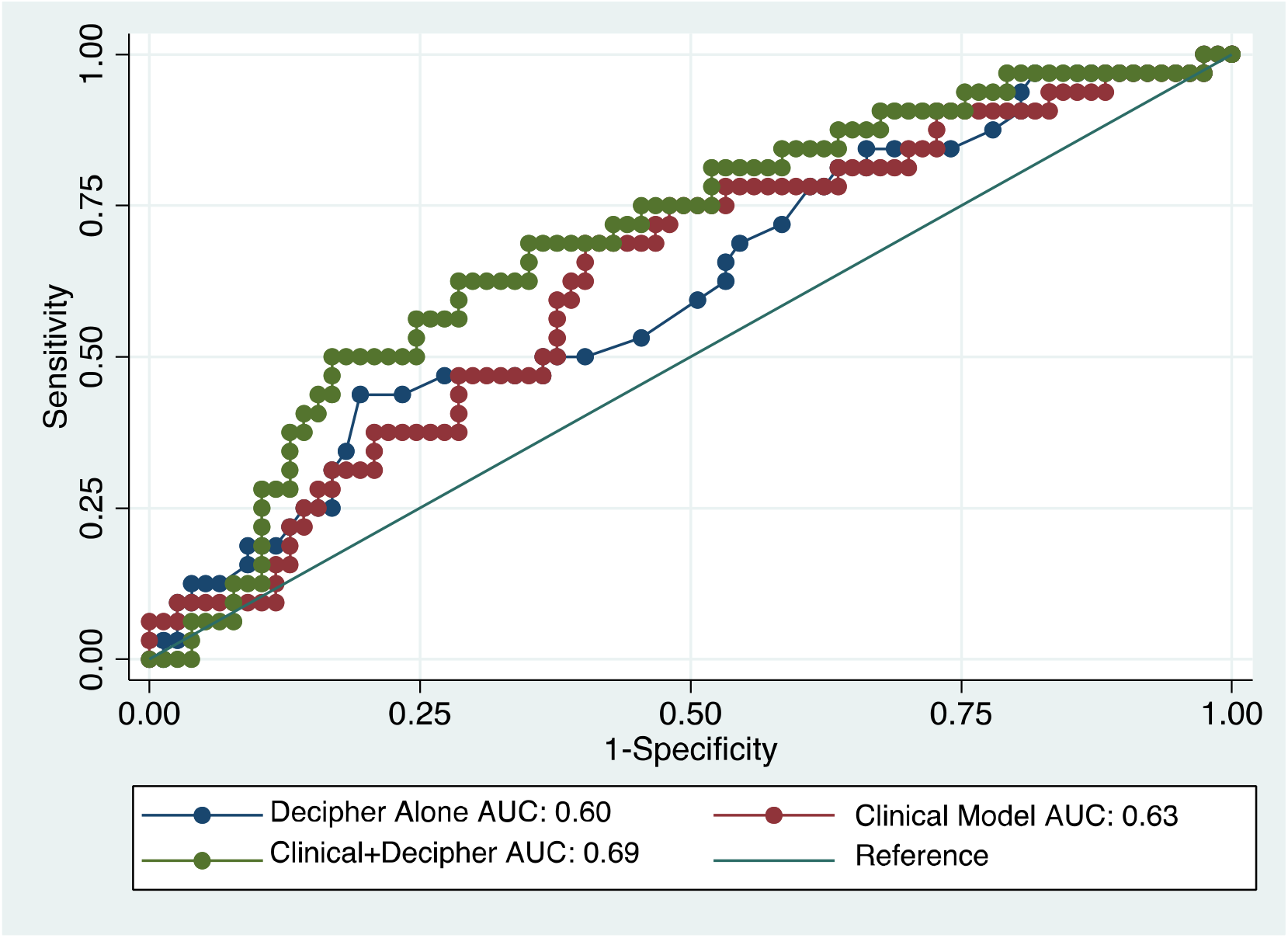
Discriminative performance of clinical models for the prediction of biopsy upgrade during active surveillance with and without the integration of Decipher.

## Discussion

In this study, we found that the Decipher genomic classifier was associated with subsequent biopsy upgrading among patients enrolled on active surveillance for low-risk or favorable-intermediate-risk prostate cancer. In this contemporary cohort of patients undergoing prostate MRI-ultrasound fusion biopsy, the Decipher score stratified the risk of reclassification independently of clinical features, including PSA density and MRI findings. Integration of the Decipher score improved the discriminative performance of a model based on baseline clinical parameters and prostate MRI, although overall performance remained modest. We further found that, based on a lower distribution of Decipher scores in the active surveillance population, reported risk groupings were not informative for estimating the probability of Gleason upgrading. As a result, distinct cut points or regard for the classifier as a continuous measure of risk may have the most utility in active surveillance. These data provide novel quantitative information for possibly integrating this baseline genomic classifier information into clinical counseling.

Among patients with GG1 but not GG2 prostate cancer electing active surveillance, Decipher scores were independently associated with Gleason upgrading on a subsequent biopsy. This additional predictive information may have greater utility in low-risk patients than low-intermediate-risk patients that have a 2 to 4-fold increased risk of reclassification, based on clinical and pathology parameters.^18,19^ Although a large body of evidence has accumulated investigating the associations between the Decipher score and clinical and pathology outcomes, there is little direct evidence concerning its short-term prognostic significance in active surveillance patients. The findings from this study suggest that the Decipher classifier may be useful in identifying patients whose initial biopsies may have been misclassified or will experience progression of their disease in the short term. However, we did not identify a significant association between Decipher score and biopsy upgrade among the subset of patients with GG2 disease. This may reflect the smaller sample size relative to GG1 and lower power for this comparison, the contributions of biopsy sampling leading by chance to the detection of a higher proportion of Gleason pattern 4 disease, or the possibility that the Decipher score is indeed not associated with further biopsy upgrade in this group.

The setting of this cohort within the contemporary era of MRI-ultrasound-fusion-guided biopsy increases the generalizability of the results, as MRI imaging is increasingly used to improve the initial assessment of cancer grade, but does not eliminate misclassification.^5^ Within this context, we found that baseline clinical paramters—including PSA density, number of cores positive for cancer, MRI findings and age—offered only marginal discriminitive ability for the prediction of biopsy upgrade but were improved by the addition of the Decipher classifier. Therefore, further optimization of prediction tools for active surveillance outcomes remains an important and still unfulfilled clinical need. ^20,21^

Our findings build upon prior studies of surrogate endpoints for active surveillance candidacy. For example, Herlemann and colleagues evaluated 647 patients diagnosed with the National Comprehensive Cancer Center Network (NCCN) with very-low, low-, and favorable-intermediate-risk prostate cancer treated with initial prostatectomy. In this cohort, the Decipher score was an independent predictor of adverse pathology, i.e., high-grade and/or high-stage at prostatectomy, (OR 1.34 per 0.1 unit increase, 95% CI 1.11-1.63).^12^ Similarly, Kim and colleagues analyzed Decipher scores from the biopsies of 266 patients with NCCN very-low, low-, and favorable-intermediate-risk prostate cancer and also reported that the Decipher score was an independent predictor of adverse pathology on prostatectomy (odds ratio 1.29 per 10% increase, 95% CI 1.03–1.61 per 0.1 unit increase).^22^ However, by directly evaluating outcomes of patients actually on active surveillance, we identified a significant association of Decipher test results and tumor upgrading. Furthermore, as reclassification events constitute the most significant triggers for conversion to active treatment, our results may have implications for questions of health-related quality of life and cost-effectiveness in future studies.

The reclassification events in our study were assessed over a relatively short interval after enrollment in active surveillance. Nearly one-third of patients in this study experienced biopsy Gleason upgrading – a larger proportion than reported from large, institutional cohorts such as the multi-institutional Canary Prostate Active Surveillance Study in which 27% of patients experienced Gleason reclassification at a median follow-up of 4.1 years.^23^ This suggests that the upgrading observed was largely due to initial biopsy sampling error rather than disease progression.^24^ On the other hand, serial molecular profiling of prostate biopsies using immunohistochemistry and next-generation sequencing has also identified the potential contributions of short-term clonal progression of low-grade disease.^25^ Regardless of the cause of upgrading, the potential role of a genomic classifier to enhance estimates of a patient’s trajectory at the time of active surveillance addresses an important clinical need. The overall modest performance of even a refined clinical model incorporating prostate MRI and genomic testing in this study underscores the need to improve risk estimation for patients enrolled on active surveillance.

We found that the distribution of Decipher scores among active surveillance patients is narrower and, as would be expected, was clustered at the lower end of the risk distribution. As a result, different groupings may be required for distinguishing risk among active surveillance patients, as the existing reporting classifications may be better suited for the wider spectrum of genomic risk. Although the Decipher score as a continuous variable (per 0.1 unit) was associated with Gleason upgrading, significant differences could not be appreciated when using Decipher’s standard risk groups generated in clinical reporting (low, intermediate, and high) that are applied in the setting of more advanced disease. Assessing a putative clinical cut point that would maximize sensitivity and specificity in this select group of patients yielded a value of 0.475, a binary classification in which values above this threshold were associated with a nearly four-fold higher odds of biopsy upgrading. A theoretical clinical application of these findings would include offering Decipher testing broadly to patients with Gleason 3+3 disease in whom identifying disease reclassification would lead to actionable differences in management, such as those with a stronger inclination for undergoing definitive treatment due to younger age, less comorbidity, and preference. For those patients bearing the lowest risk of disease reclassification, as assessed through clinical and genomic features such as a Decipher score <0.475, reducing the intensity of surveillance by increasing intervals between biopsy, or avoiding biopsy altogether, may be feasible. However, these findings require further study in larger cohorts and over longer periods of surveillance, or explicit study in a randomized trial.

There are several limitations of this study. Selection of patients for Decipher testing may not have occurred at random and could potentially favor use in patients at higher risk for disease reclassification for whom testing was undertaken to confirm suitability for active surveillance. However, Decipher testing was routinely offered without known systematic preference for those at higher risk, and patient baseline disease characteristics are consistent with widely-accepted criteria for adoption of active surveillance.^6^ A higher incidence of short-term reclassification also was reported in prior studies of patients receiving genomic testing that may relate to the preferential use of genomic testing in higher-risk populations.^26^ In addition, we defined biopsy upgrade as any increase in biopsy Gleason score, an approach used in prior studies, but this may fail to account for more substantial changes of risk such as a simultaneous increase in tumor volume.^27^ Although all prostate MRI studies were reviewed by expert genitourinary radiologists at our institution, a central re-review of studies was not conducted for this study to apply the PRECISE criteria for MRI progression.^28^ Lastly, there is an insufficient sample size and follow-up to assess the meaningful distant longitudinal outcomes. Despite these limitations, strengths of this study include novel data on Decipher testing with outcomes of patients enrolled in a contemporary active surveillance program.

## Conclusion

The Decipher genomic classifier score was associated with biopsy Gleason upgrading among patients with low-risk prostate cancer enrolled in active surveillance who had undergone MRI-enhanced biopsy procedures.

## Data Availability

All data produced in the present study are available upon reasonable request to the authors

